# Low-cost generation of clinical-grade layperson-friendly pharmacogenetic passports using oligonucleotide arrays

**DOI:** 10.1101/2024.08.28.24312707

**Authors:** Pauline Lanting, Robert Warmerdam, Jelle Slager, Harm Brugge, Taichi Ochi, Marloes Benjamins, Esteban Lopera-Maya, Soesma Jankipersadsing, Jody Gelderloos-Arends, Daphne Teuben, Dennis Hendriksen, Bart Charbon, Lennart Johansson, Thijs Oude Munnink, Nienke de Boer-Veger, Lifelines NEXT, LifeLines Cohort Study, Bob Wilffert, Morris Swertz, Daan Touw, Patrick Deelen, Nine Knoers, Jackie Dekens, Lude Franke

## Abstract

Pharmacogenomic (PGx) information is essential for precision medicine, enabling drug prescriptions to be personalized according to an individual’s genetic background. Almost all individuals will carry a genetic marker that affects their drug response, so the ideal drug prescription for these individuals will differ from the population-level guidelines. Currently, PGx information is often not available at first prescription, reducing its effectiveness. Pharmacogenetic information is most often obtained using special assays, making it expensive and time-consuming to generate. We therefore hypothesized that we could also use genome-wide oligonucleotide genotyping arrays to generate comprehensive PGx information (PGx passports), thereby decreasing the cost and time required for PGx testing, and lowering the barrier to generating PGx information prior to first prescription.

Taking advantage of existing genetic data generated in two biobanks, we developed and validated Asterix, a low-cost clinical-grade PGx passport pipeline for 12 PGx genes. In these biobanks we performed and clinically validated genetic variant calling and statistical phasing and imputation. In addition, we developed and validated a novel *CYP2D6* copy number variant calling tool, foregoing the need to use separate PCR-based copy number detection. Ultimately, we returned 1227 PGx passports to biobank participants via a layperson-friendly app, improving knowledge of PGx among citizens. Our study demonstrates the feasibility of a low-cost clinical-grade PGx passport pipeline that could be readily implemented in clinical settings to enhance personalized healthcare, ensuring that patients receive the most effective and safe drug therapy based on their unique genetic makeup.

## 1. Introduction

There is now extensive knowledge available on pharmacogenes, and clinical guidelines are available for personalized drug treatment using genetic data (1). However, the application of pharmacogenetics (PGx) in clinical practice remains limited (2) and is mostly done reactively to understand why patients reacted poorly to prescribed drugs. However, a pre-emptive pharmacogenetic passport would allow results to be stored in (electronic) medical files for future medical use (3), ensuring that PGx data are available at first prescription, thereby enabling optimized treatment and preventing adverse effects (4,5). Currently, the major impediments to such pre-emptive testing are cost and speed: PGx information is most commonly obtained using dedicated assays that target a limited subset of haplotypes of a single gene. This makes the generation of a complete, multi-gene profile time-consuming and costly.

In theory, PGx data can also be obtained from genetic data that is routinely used in both research and clinical genetics. Single nucleotide polymorphism (SNP) microarrays in particular have proven essential for detection of Copy Number Variations (CNVs) in clinical genetics and genome-wide association studies (GWAS) in research (6,7). However, whether this data can be used to provide a clinical-grade PGx passport to individuals is largely unknown, and there are a number of challenges that need to be resolved before this data can be effectively used for PGx. Firstly, SNP arrays do not consider every variant and do not capture on which haplotype specific information is located, requiring the use of genotype phasing and imputation. Secondly, *CYP2D6*, an important gene in pharmacogenomics (8) is highly polymorphic, and it is also affected by CNVs, making the genotyping of this gene non-trivial. To deliver on the promise of personalized medicine, we need to bridge these gaps in order to translate available genetic data to clinically relevant PGx data.

In parallel with the decreasing cost of genetic testing over the past decade, the number of large-scale biobank initiatives has also increased. Jointly, these biobanks have now generated genotype data for millions of individuals (9–12). This provides an opportunity to repurpose readily available genetic data to develop and validate the use of SNP array data for generating PGx passports. This would allow researchers to return clinical-grade PGx results to biobank participants, thereby decreasing the barriers to applying pre-emptive PGx testing and facilitating personalized medicine in clinical settings.

Another problem is that the disclosure of PGx information so far has mainly been limited to researchers, clinicians, and pharmacists. Yet sharing relevant PGx information with citizens is important to improve knowledge and understanding of PGx testing and its related health outcomes. Digital health technologies, including mobile phone applications, have increased citizen engagement in the last 10 years (13,14), and they allow patients to be engaged in the healthcare decision-making process. Implementation of digital health technology in optimizing medication adherence in patients with depression, respiratory diseases, and cancers has been shown to improve patient outcomes. However, the use of digital health technologies to provide PGx information to patients has been limited. However, with growing populations throughout the world, governments are now actively stimulating the development and use of such tools. For instance, the recent Integral Care Agreement in the Netherlands highlights the importance of digitalization to promote self-reliant digital healthcare (15).

In the northern Netherlands, the Lifelines Cohort Study, a multigenerational, longitudinal, dynamic cohort and biobank, presents a unique opportunity to leverage available genetic data to facilitate healthy aging (16). This biobank has been genotyping its participants for multiple years, as has the separate Lifelines NEXT prospective birth cohort biobank. In this study, we developed and clinically validated 1) an automated software tool for the interpretation of array-based genotype data to generate reliable PGx profiles, 2) a layperson-friendly translation of Dutch Pharmacogenetics Working Group (DPWG) guidelines, and 3) a digital infrastructure to communicate these to biobank participants. By developing this tool, we aim to improve the lives of patients and the general population, starting with the population of the northern Netherlands and expanding to more individuals in the future.

## 2. Methods

### 2.1 Datasets and quality control

We used four different datasets to develop and validate the automated generation of PGx passports, divided over two different versions of Illumina Genotyping arrays. We used 38,030 samples from the UMCG Genetics Lifelines Initiative (UGLI) cohort of the Lifelines prospective follow-up biobank (17) to develop and validate an automated PGx passport pipeline using the Infinium Global Screening Array® MultiEthnic Disease (GSA-MD) version 1.0 array. In parallel, we used three datasets to develop the pipeline for the GSA-MD version 3.0 array: 2473 samples from our Genome Diagnostics patient cohort (UMCG), 77 validation samples with known PGx genotypes, and another 574 samples from the Lifelines NEXT cohort (18).

The samples of the Lifelines UGLI cohort were genotyped using GSA-MD version 1.0. We performed quality control on both samples and markers, including removal of samples and variants with a low genotyping call rate (<97%), variants showing deviation from Hardy-Weinberg equilibrium (p < 1×10^−6^), excess of Mendelian errors in families (>1% of the parent–offspring pairs), and samples with very high or low heterozygosity (4 standard deviations (SD) above or below the mean for autosomes). We further revised and removed samples that did not show consistent information between reported sex and genotypes on the X chromosome, or between reported familial information and observed identity-by-descent sharing with family members. All other samples were genotyped using GSA-MD version 3.0 and were quality-controlled similarly to those in Lifelines UGLI. In total, 36,373 Lifelines UGLI samples, 470 Lifelines NEXT samples, 2285 validation samples of the Genome Diagnostics patient cohort, and 76 validation samples with known PGx genotypes passed quality control.

### 2.2 Selecting individuals to receive pharmacogenetic results

Quality control measures in research settings are commonly less stringent than in diagnostics. Since Lifelines UGLI samples were genotyped in a research setting, this dataset could potentially contain sample swaps. Therefore, we used Idéfix (19) to select a high-quality subset of 5,765 samples suitable for returning personalized results. In short, Idéfix selects those samples for which the actual phenotypes sufficiently match the predicted phenotypes based on genetic predispositions. These genetic predispositions were determined by calculating polygenic scores (PGSs) per phenotype. These calculations aggregate the effects of common risk-decreasing or -increasing alleles identified by GWAS. Here, we reran Idéfix using an updated list of PGSs based on recent GWAS. Quality control, PGS calculation, and running Idéfix were performed as described in Warmerdam et al. (19). Finally, we returned PGx results to 996 active Lifelines participants who gave consent to receive their PGx passport. For Lifelines NEXT, we were able to manually resolve sample swaps based on family data and duplicated array runs. We returned PGx results to 231 mothers registered as active Lifelines NEXT participants who gave consent to receive their PGx passport.

### 2.3 Variant calling

Variants were called in two separate stages. In general, we used the Opticall software (20) to do genotype-calling on the majority of variants. However, Opticall is not suitable for genotyping variants covering frequent CNVs. Since CYP2D6 is affected by relatively frequent CNVs, it is not trivial to genotype variants within and near this gene. At the same time, inclusion of CYP2D6 is essential to return a comprehensive PGx passport because it metabolizes a large proportion of medication. We therefore developed a novel algorithm for CNV-calling of the CYP2D6 gene using the data obtained from a SNP array. In addition, this method also performs genotype-calling of individual variants in the CYP2D6 gene.

#### 2.3.1 CYP2D6

SNP arrays have been used extensively to study both SNPs and CNVs. Modern SNP arrays genotype at hundreds of thousands of assays (probes designed to genotype one particular variant) simultaneously by measuring the color and intensity of light emitted when one of four labeled nucleotides hybridizes at the assays. In addition to enabling study of which alleles are present in an individual through comparison of the colors of the emitted light, it also allows study of CNVs by combining the total signal intensity. We combined these two approaches in our *CYP2D6* genotyping methodology, as described below.

For genotyping *CYP2D6*, we first selected a number of independent, high-quality variants from the quality-controlled genetic data (missingness < 0.01, minor allele frequency (MAF) > 0.05, Hardy-Weinberg equilibrium p-value > 0.05, *HLA* excluded, minimum distance of 100kb between variants, R^2^ < 0.4). Using these variants, our method performs a principal component analysis of the intensities of the selected variants (summed across the two alleles). As the first principal components (PCs) likely tag batch effects, we chose to regress out the first 100 PCs from the genomic region of interest – the *CYP2D6* gene – using ordinary linear regression (OLS). Both PC loadings and OLS regression parameters can be estimated in one dataset, and later applied in another. The methodology for correcting SNP array intensities follows the approach previously established by Lude Franke et al. (21).

After correction for batch effects, one variant is selected to assign naive CNV-calls (rs1135824 for GSA-MDv1, rs1423323203 for GSA-MDv3). Naive, preliminary CNV assignments are made by binning the total signal intensity of the selected variant so that the proportions of CNV genotypes match the expected frequency of CNV-calls. The expected frequencies (*5 = 0.0295, Duplication = 0.0275) are obtained from PharmGKB (https://www.pharmgkb.org/page/cyp2d6RefMaterials, European population). Using the means and SDs of the signal intensities for each bin, a naive, preliminary CNV call is assigned to every individual based on which CNV bin has the highest likelihood at the signal intensity of the specific variant. If multiple GSA probes are available for a variant, a majority voting ensemble classifier is used to obtain a consensus of the various assays. Next, genotyping is performed using a manually selected set of variants. These variants are selected based on whether the individual signal intensities show clusters of genotypes in accordance with what can be expected for a biallelic variant with possible deletions and duplications (i.e., at least six visible intensity clusters rather than the three intensity clusters expected when dealing with a biallelic variant outside a CNV).

For each variant and possible copy number, a nearest-neighbors classifier is used to obtain a set of highly confident samples for every variant. Then, to perform genotyping, the theta of every sample is calculated based on its intensities. For this, the median of the intensities for the samples naively assigned to the group of homozygous deletions is used as the origin. Thereafter, samples are binned into k bins, where k corresponds to the number of genotypes that can be expected for a biallelic variant at each copy number. Bins are assigned genotypes so that they best match Hardy-Weinberg principles. Per variant, naive genotype clusters are then iteratively improved in a gaussian mixture model algorithm adapted from *scikit-learn* (22). The maximization step in the gaussian mixture model algorithm was changed to also weight genotypes according to Hardy-Weinberg principles, which were expanded to also take deletions and duplications into account. Per variant, this procedure is performed while accounting for the intensity measures of all assays tagging that specific variant.

For each combination of a variant, sample and copy number, a total probability is calculated by summing the probabilities across the genotypes corresponding to that copy number. Subsequently, the most likely copy number is assigned to each variant-sample combination by selecting the copy number with the largest total probability among the possible copy numbers. The CNV status of *CYP2D6* as a whole is determined by summing the probabilities of variants over all possible copy numbers, and rescaling these to sum to one. Genotype probabilities are then calculated by rescaling the possible genotype probabilities in the assigned copy number status to one.

Genotypes of samples with 1 or 2 *CYP2D6* copies (across both the maternal and paternal chromosome) are written so that genotypes of samples with single-gene deletions are homozygous for the reanalyzed variants. Genotypes for variants of samples that are homozygous for whole-gene deletions do not exist. In human genetic datasets, the representation of genotypes for variants of samples with a *CYP2D6* duplication is not trivial, making the imputation and phasing non-trivial. Therefore, these samples are excluded. *CYP2D6* variants in chromosome 22 genotyped using Opticall are replaced with the newly genotyped *CYP2D6* variants. This is subsequently used in the phasing and imputation pipeline. The *CYP2D6* CNV-calling algorithm is included as the first part of the *Asterix* software that we developed to generate PGx profiles.

### 2.4 Phasing and imputation pipeline

Before phasing and imputation, PLINK files are lifted from Human genome build 37 to build 38 and alleles of insertions and deletions are recoded to match the notation in the reference dataset. Then, variants for which the strands do not match the reference panel are removed, and variants with >5% missingness or a Hardy-Weinberg p-value < 1×10^-6^ are removed. Phasing and imputation are implemented using Eagle v2.4.1 (23) and Minimac4 (24)(version 1.0.2). The reference dataset for phasing and imputation is derived from the 1000 Genomes project 30x coverage whole genome sequencing dataset (25). Finally, we replaced the imputed genotypes with the original genotypes for the variants that were used as input in the imputation.

### 2.5 Crucial backbone

A minimum set of variants that should be included in our genotyping panel was determined per gene (see Table S1). We used available ‘tier 1’ recommendations for *CYP2C9*, *CYP2C19*, *CYP2D6, CYP3A4, CYP3A5,* and *VKORC1* (26–29). For the other genes in our PGx passport, we assessed the variants/alleles that were mentioned in DPWG background information and those that are included in our validation dataset. We included a variant in the crucial backbone when: 1) the variant results in an allele known to alter the drug response phenotype, and 2) the MAF in the (North-Western) European population is > 0.001.

### 2.6 Validation

We validated GSA-MD versions 1 and 3 and *Asterix* following the clinical genome diagnostic standards in our center. The validation parameters and corresponding acceptance criteria are shown in Table 1.

**Table 1:** Validation parameters and acceptance criteria for the clinical validation of the Infinium Global Screening Array MultiEthnic Disease versions 1 and 3, and the software tool Asterix. ‡For all variants available in both gnomAD-NFE and GoNL datasets, the distribution of mutant allele frequency differences between those two datasets was recorded. Z represents the standard deviation of that distribution. Variants that deviated more than 3*Z from the gnomAD-NFE mutant allele frequency failed validation.

| Topic | Validation parameter | Acceptance criterium |
| --- | --- | --- |
| Quality of the array data | Call rate | <sup>3</sup>0.97 |
| Phasing and imputation quality | Concordance with trio-data | <sup>3</sup>0.97 |
|  | R<sup>2</sup> | <sup>3</sup>0.97 |
| Result interpretation | Absolute number of mutant alleles detected | <sup>3</sup>5 |
|  | Mutant allele frequency | <3*Z<sup>‡</sup> |
|  | Concordance with known genotypes | 100% |

We created a benchmark set of haplotypes using data of 621 (GSA-MD v1) and 135 (GSA-MD v3) parent–offspring trios to validate phasing performance. Using *SHAPEIT5* (30) we then compared the results of our statistical phasing with the benchmark haplotypes and computed the switch error rates (SER) of all genetic variants that were directly typed. The call rate and imputation quality are validated based on the output of our quality control procedures. Variants were removed if: 1) they were not directly genotyped nor imputed, 2) they were not in the crucial backbone and had a low imputation probability (R^2^ < 0.97), 3) the mutant allele was observed < 5 times in the full dataset, 4) the mutant allele frequency deviated more than 3*Z from the gnomAD-non-Finnish Europeans (NFE) mutant allele frequency (Z represents the SD of differences in mutant allele frequency between the GoNL and gnomAD-NFE datasets), 5) the phasing SER was > 0%, or 6) the concordance with known genotypes of validation samples was < 100%.

When a variant was in the crucial backbone, but the imputation probability was low, we used haploid-level dosage (HDS) to assess the variant call confidence level in an individual sample.

In addition to variant calling, we also validated our CNV-calling algorithm. To that end, we selected samples with a confidence score of > 0.97 (1x zero copies, 14x one copy, and 16x two copies) and samples with a confidence score of with a CNV-calling confidence score of > 0.9, and < 0.97 (4x one copy, 3x two copies and 5x three copies). DNA of these samples was analyzed with a dedicated TaqMan real-time PCR assay specifically targeting exon 9 of the *CYP2D6* gene (Certe, Groningen, the Netherlands). It is worth noting that this assay cannot detect gene duplication or hybrid alleles. However, as we report *CYP2D6* to be not available when >2 copies of *CYP2D6* are detected, these calls do not currently require validation.

Finally, we confirmed that the software we developed correctly translated 1) the detected mutations to the corresponding star alleles and, subsequently, 2) the combination of haplotypes to the corresponding contraindication (CI) and/or predicted phenotype. This is described in more detail in section 2.9.3 and in the Supplementary Information.

### 2.7 Pruning of allele definition tables

We used gene-specific allele definition tables to assign star alleles. Available tables were downloaded from PharmVar (v5.2.14 for GSA-MD version 3; v6.0.5 for GSA-MD version 1) for *CYP2B6, CYP2C9, CYP2C19, CYP2D6, CYP3A4, CYP3A5* and *NUDT15* and from PharmGKB for *ABCG2* and *TPMT* (https://www.pharmgkb.org/page/pgxGeneRef). Custom tables were constructed for *MTHFR, SLCO1B1* and *VKORC1*. We curated all allele definition tables for the GSA-MD versions 1 and 3 separately and removed all variants that did not pass acceptance criteria (Tables S2a-b). We then pruned each table to prevent ambiguous calls caused by haplotypes that we cannot distinguish based on the variants included in our pipeline. The resulting allele definition tables are shown in Tables S3a-x.

### 2.8 Translation to pharmacogenetic contraindication

To translate the diplotype to the PGx CI used in medication surveillance, we designed custom translation tables (see Tables S4a-l). Allele function of individual star alleles and their translation into predicted phenotype and PGx CI are based on PharmVar, PharmGKB, and/or DPWG background information. All possible diplotypes based on the pruned allele definition tables are included in the translation tables. We made sure the PGx CIs correspond to those used in the DPWG recommendations.

### 2.9 Generating PGx profiles

We developed the *Asterix* tool to produce PGx profiles from genotyping data from the GSA-MD versions 1 and 3. *Asterix* is composed of four main tasks: 1) determining the *CYP2D6* copy number status, 2) determining star alleles, 3) translating these to predicted phenotypes and PGx CIs, and 4) constructing a logging file.

#### 2.9.1 Determining star alleles, predicted phenotypes, and PGx CIs

After correction of the *CYP2D6* genotypes based on the CNV status, star alleles for all included genes are assigned using the gene-specific allele definition tables.

Subsequently, predicted phenotypes and PGx CIs are determined with the custom translation tables. When determining star alleles present for *CYP2D6*, *Asterix* takes into account whether whole-gene deletions (*CYP2D6*5*) were detected in CNV-calling.

#### 2.9.2 Constructing a logging file

To be able to check *Asterix* output, an extensive logging file is constructed that contains information for several checkpoints in our pipeline. Parameters that are logged include the estimated phased haploid-level dosage (HDS) when < 0.97 (only for imputed genotypes), missing variants, and haplotypes that were not described in translation tables. Identical haplotypes are merged together to simplify the validation process.

#### 2.9.3 Approval and release of the PGx profiles

The PGx profiles generated are to be released by a registered pharmacist. To maximize process efficiency, the approval and release of the PGx profiles is performed on the level of unique combinations of detected mutations and unreliable calls. When a sample contains only previously approved haplotypes, it is automatically released. Each decision is logged to maintain an audit trail. The procedure for approving and releasing the profiles is described in more detail in the Supplementary Information.

### 2.10 Building layperson-friendly PGx passports

The output of *Asterix* is a PGx profile that is suitable for application in clinical practice by healthcare practitioners. However, (future) patients should not be expected to understand the meaning or implications of such a profile. We therefore developed a layperson-friendly digital PGx passport to fulfill the informational needs of (future) patients.

#### 2.10.1 Developing layperson-friendly communication materials

We aimed to develop communication materials that explain the impact of drug–gene interactions (DGIs) that are understandable for 90% of the study population. We developed layperson-friendly texts based on the DPWG guidelines for healthcare practitioners (see Table S5), written at Common European Framework of Reference Common Reference Level B2 (31). One important difference that we had to address is that the DPWG guidelines do not contain an explanation for normal metabolizers because standard-of-care applies for these patients. However, texts for the (future) patient do need to cover this topic because it is valuable to explain that standard-of-care applies. The texts are constructed using three building blocks (Figure 1).

**Figure 1:**
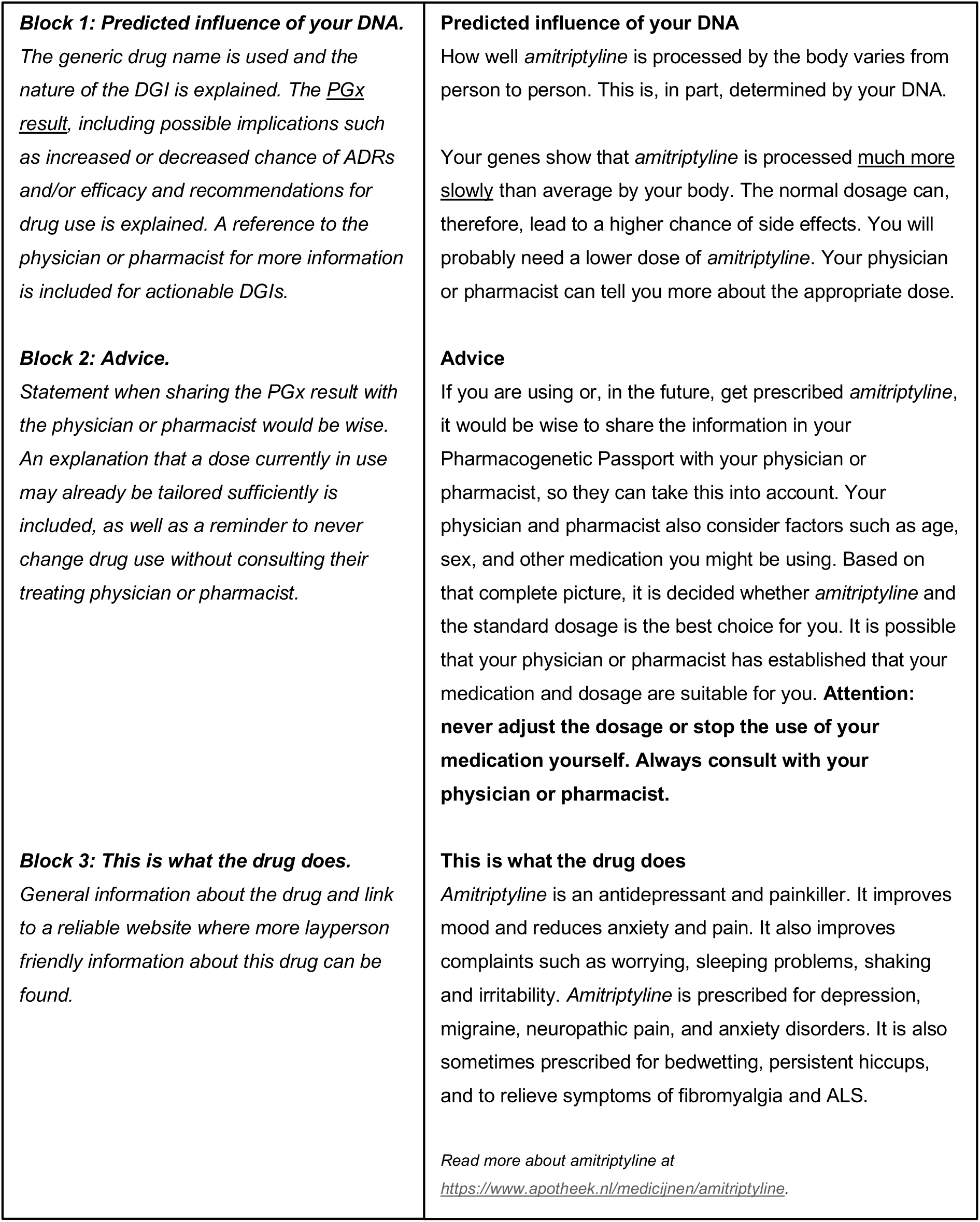
Building blocks used to construct layperson-friendly texts to explain Drug–Gene interactions (left) and the corresponding text for CYP2D6 PM-amitriptyline (translated from Dutch, right).

#### 2.10.2 Digital infrastructure

We created a DGI table that contains all the CIs produced by *Asterix*, a list of known DGIs currently in the DPWG guidelines, and the layperson-friendly texts associated to each combination of these. The relevant texts for each participant are selected based on their PGx profile and known DGIs. For example, the text explaining the consequences for amitriptyline in Figure 1 will be selected for the individuals who are listed as CYP2D6 PM in their PGx profile. The PGx profile and corresponding texts are subsequently unlocked through an Application Programming Interface (API) following the Health Level Seven Fast Healthcare Interoperability Resources (HL7-FHIR) format. Participants can view their PGx passport in the mobile application (app) *Gen en Geneesmiddel,* which we developed with Synappz Digital Healthcare and which is available for iOS and Android (see Figure 5).

#### 2.10.3 Additional features

Upon installing the app, the user is shown a series of information screens that explain PGx and the content and use of the app. This information can be reviewed within the app after completion of this initial onboarding process. Furthermore, frequently asked questions (FAQ) are included in the app to address several anticipated questions. The FAQ-section is supplemented by new questions from users and associated answers.

The layperson-friendly texts displayed in the mobile application are accompanied by articles that describe the PGx CI that interacts with the drug being viewed (see Table S5). When multiple PGx CIs are relevant for the drug being viewed, all the relevant articles are shown. The articles provide interested participants with more detailed information and facilitate communication between (future) patients and healthcare practitioners.

Finally, a technical report aimed at healthcare practitioners can be downloaded from the app. This report contains the PGx profile and accompanying information, including contact information for a helpdesk and instructions on how to interpret and process the results.

## 3. Results

### 3.1. Pharmacogenomic profiles and passports

To validate the use of a layperson-friendly, clinical-grade PGx passport using oligonucleotide arrays, we have developed a tool (*Asterix*) to produce PGx profiles from genotyping data from the GSA-MD versions 1 and 3. As described in Figure 2a, our tool works by determining the *CYP2D6* copy number, phasing and imputing genetic variants, determining the star alleles of the samples provided, and translating diplotypes to PGx CIs. Instructions on the installation and use of *Asterix* are described on the GitHub repository (https://github.com/molgenis/asterix).

**Figure 2:**
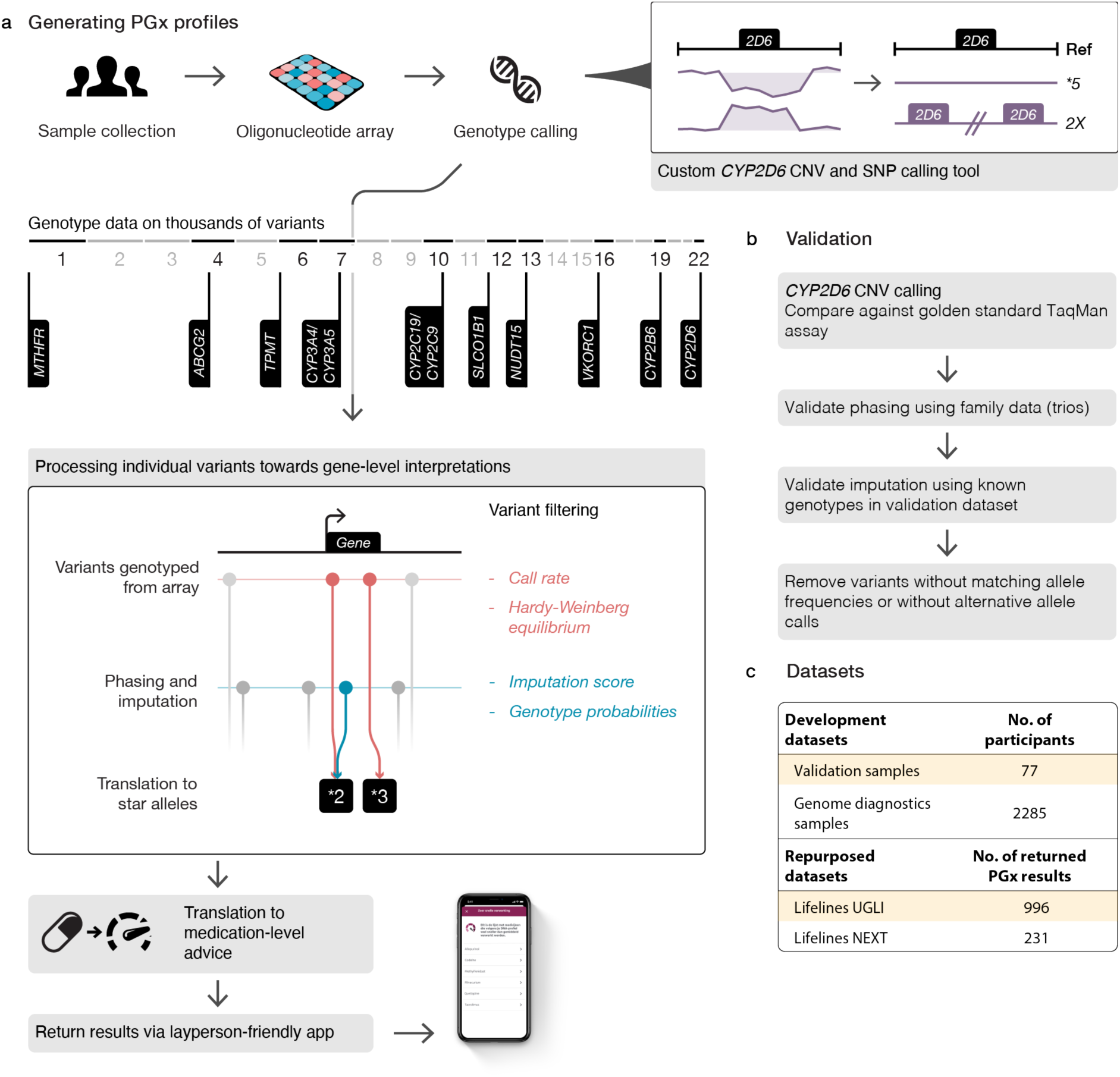
Overview of the work in this paper. (a) Schematic of the pharmacogenetic (PGx) pipeline developed to generate PGx profiles. Data collection involves obtaining DNA samples from participants and genotyping using an oligonucleotide array. We ascertain CYP2D6 copy numbers (CN) using a custom CNV calling approach. This method estimates CN status at multiple points within the CYP2D6 gene and integrates these estimates to determine the overall gene copy number status. Twelve genes contribute to the generation of PGx profiles. The combined effects of these genes are aggregated into medication-level recommendations, which are communicated to participants via a layperson-friendly app. (b) Broad overview of the methodology used for validating the PGx pipeline. (c) Overview of the datasets used for development and validation of the PGx passport (Development datasets), and those for which we also returned PGx-profiles (Repurposed datasets).

### 3.2. Validation

A total of 34 variants, in 12 genes, were included in the crucial backbone of the PGx pipeline, and subsequently underwent validation (Table 2). For *CYP2B6* we initially also included rs3211371 (*\*5*). However, since the DPWG withdrew CI definitions differentiating *CYP2B6*5* from *\*1*, we removed this SNP from the crucial backbone. Three variants in *CYP2C9* (rs28371686 (*\*5*), rs9332131 (*\*6*) and rs7900194 (*\*8*)) were initially included in the crucial backbone based on their multiethnic allele frequency (26). However, since each of these variants had a MAF < 0.001 in the NFE population, we subsequently excluded them from the crucial backbone for GSA-MD v1. It is important to note that the Lifelines population is known to be reflective of the NFE population. For the same reason, we excluded rs41303343 (*CYP3A5*7*). For *TPMT* variant rs1800462 (*\*3A*/*\*3B*), no mutant alleles were detected on GSA-MD v3. As we could not ensure the associated GSA probe was functional, *TPMT* was excluded from PGx profiles generated with GSA-MD v3. All other variants in the crucial backbone passed validation criteria. In case of an imputation score (R^2^) < 0.97, imputation quality was judged from sample-specific HDS. When the HDS < 0.97, a registered pharmacist decided during approval and release whether a result could be released for the associated gene.

**Table 2:**
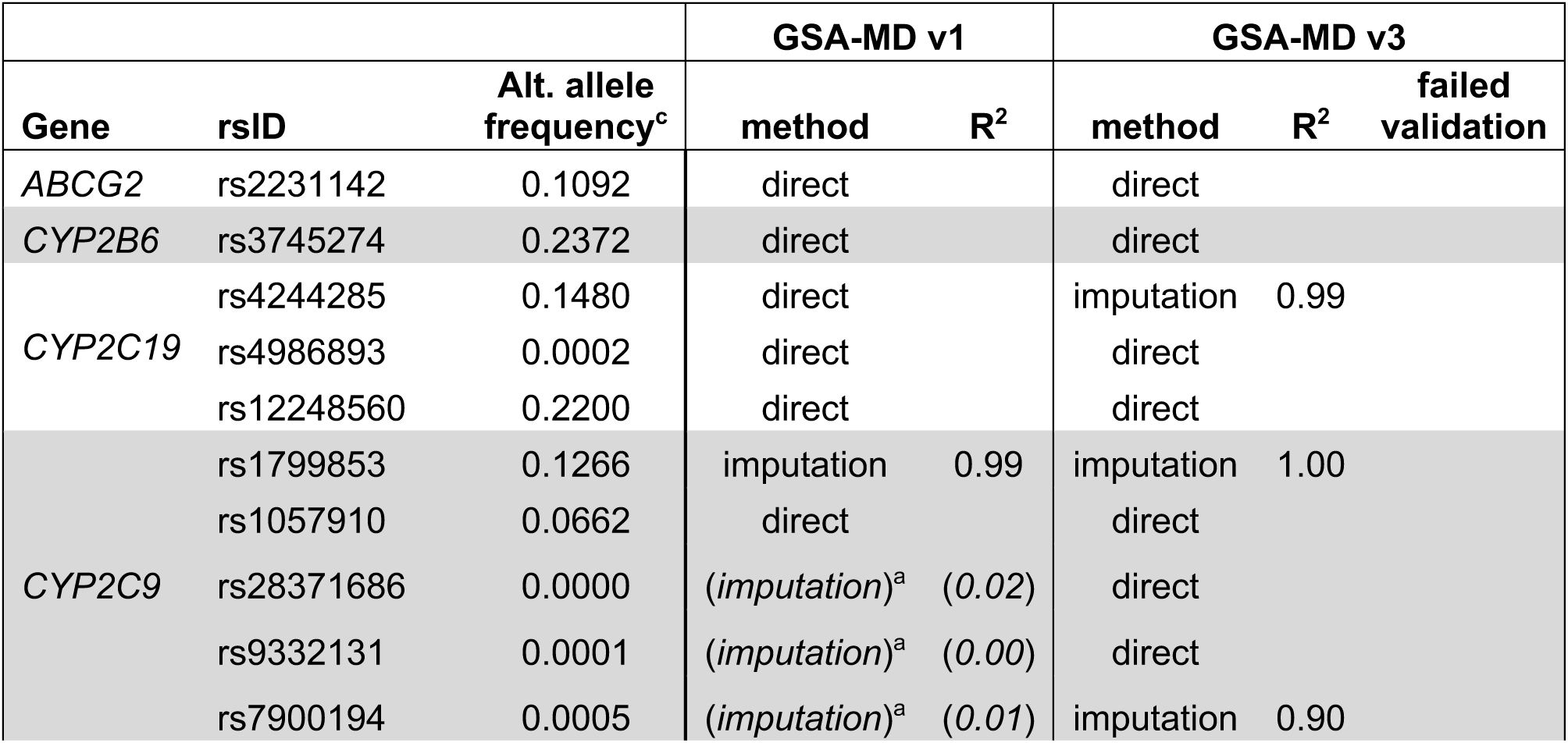
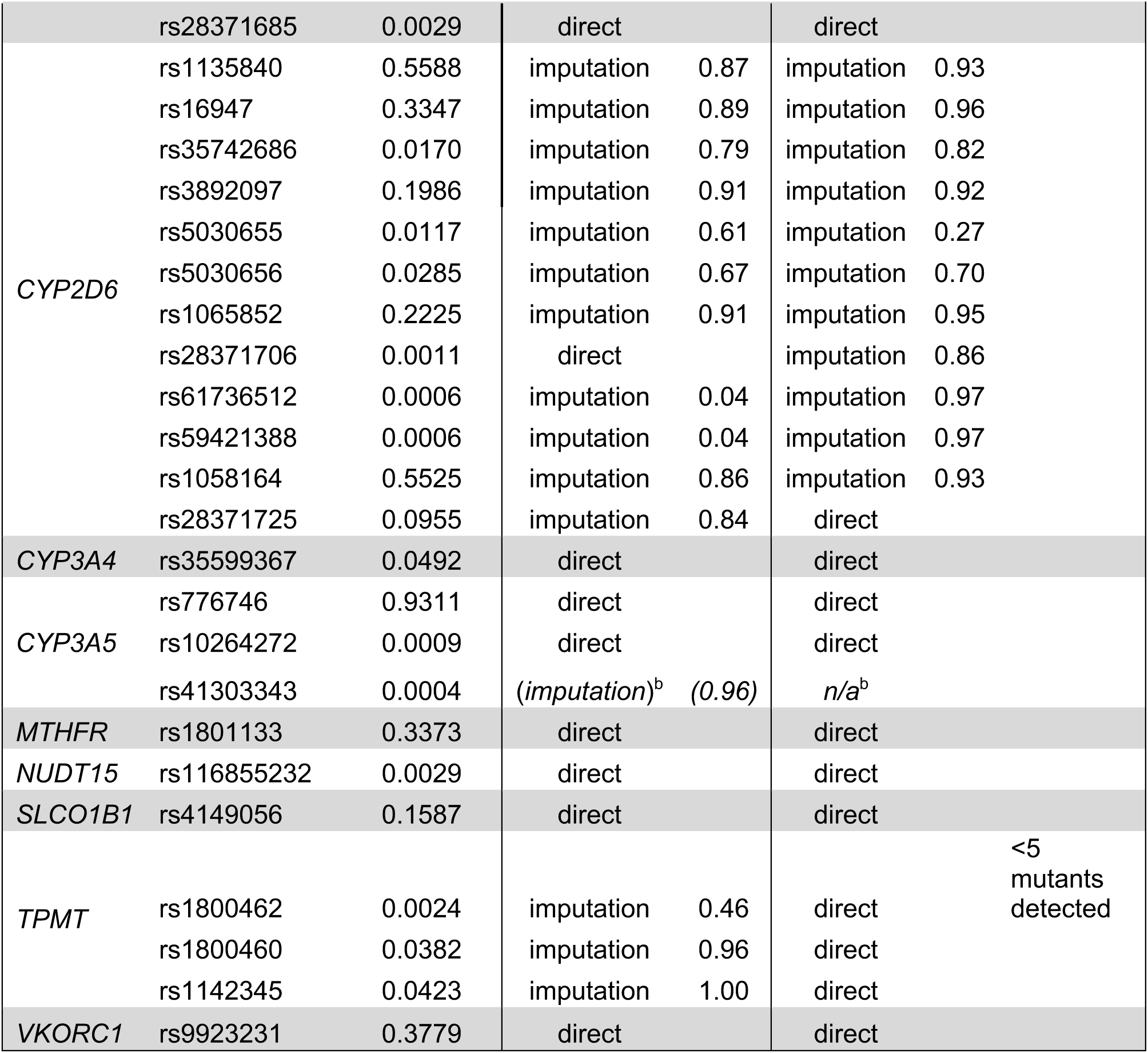
Validation results of crucial backbone variants. ^a^rs28371686, rs9332131, and rs7900194 were left out of crucial backbone for GSA-MD v1, based on known ethnic composition of Lifelines ^b^rs41303343 was left out of the crucial backbone for both GSA-MD v1 and GSA-MD v3, based on known ethnic composition of Lifelines ^c^Reference allele frequency in non-Finnish Europeans (NFE)

We considered 731 and 703 variants outside the crucial backbone for inclusion in the translation tables of GSA-MD v1 (PharmVar tables v6.0.5) and GSA-MD v3 (PharmVar tables v5.2.14), respectively. Results of the validation process are summarized in Figure 3 and are fully available in Tables S2a-b. Many variants were removed due to unavailability on the genotyping arrays and in the imputation panel. Beyond that, poor imputation score and/or low allele frequency were the leading reasons for exclusion. Phasing quality was verified by comparison to phasing results using parent–offspring trios. Poorly phased variants were retained when they were the only variant of interest in that gene, rendering phasing irrelevant. Importantly, 37 variants were typed in 76 validation samples, using a traditional DNA amplification genotyping method (Leiden University Medical Center). For all 37 variants, the genotypes derived from GSA-MD v3 (19 directly typed, 18 imputed) were a 100% match to the traditional assays.

**Figure 3:**
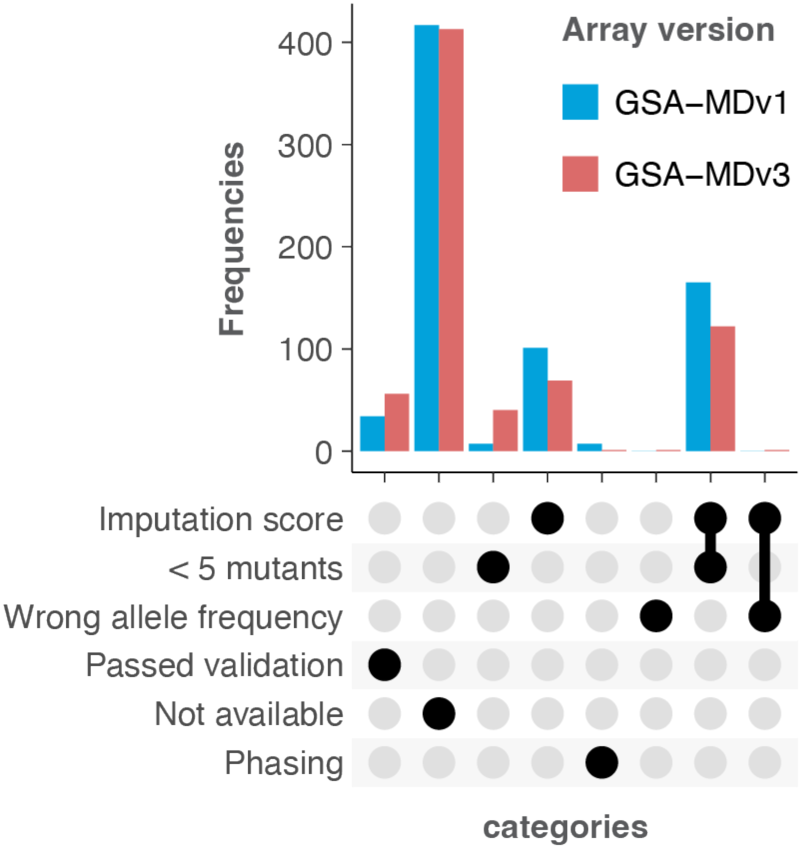
Validation status of variants from the PharmVar translation tables outside the crucial backbone. The UpSet plot displays the frequency of variants that either meet or fail to meet the validation criteria. Columns with a single marked circle represent the frequency of variants failing exclusively that criterion, while columns with multiple marked circles indicate variants that failed multiple corresponding criteria.

Within *Asterix*, we embedded a custom *CYP2D6* copy number (CN) calling algorithm. To validate the *Asterix*-derived *CYP2D6* CN calls, a total of 43 samples were selected, representing a CN probability of at least 0.9 and each of the four possible CN calls (zero copies, one copy, two copies, and three copies). The CN status of the aforementioned samples was determined using a dedicated TaqMan assay targeting exon 9. To compare the results, we initially evaluated whether the overall CN calls with a moderate probability of over 0.9 matched the calls reported by the TaqMan assay (Figure 4a). Of the 43 samples, *Asterix* identified three as having a CYP2D6 duplication (three copies), whereas the TaqMan assay reported these samples as homozygous reference (two copies). In one instance, *Asterix* reported a heterozygous deletion (one copy) while the TaqMan assay returned a homozygous reference call. However, for these four samples, *Asterix* consistently reports a homozygous reference call at exon 9, which is consistent with the call reported by the TaqMan assay. For the majority of the other variants, *Asterix* reports calls matching that of the overall aggregated estimate (Figure 4b). This suggests that these samples are likely to possess a hybrid allele or tandem arrangement, resulting in a distinct CN call at exon 9. It is worth noting that the TaqMan assay is not capable of differentiating between regular copy numbers and hybrid alleles. At a probability of at least 0.97, all 31 samples exhibit matching CN calls.

**Figure 4:**
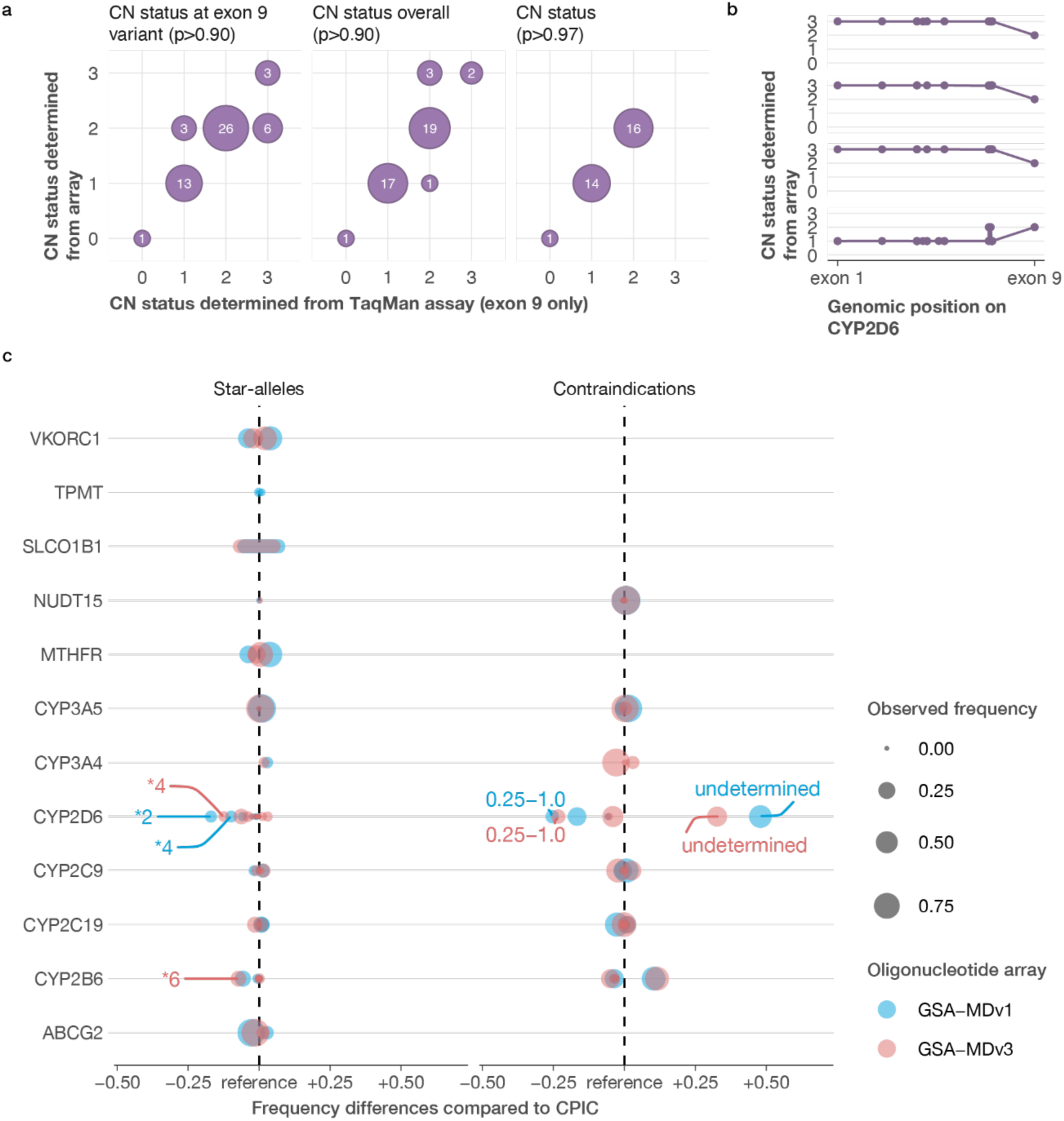
Validation results of CYP2D6 CNV-calling (a, b). a) correspondence between a standard CYP2D6 CNV-calling procedure using a TaqMan assay (x-axis) and the CNV-calling results determined using GSA-MD v3 and our specially designed algorithm. Only exon 9 was assessed in the TaqMan assay. The y-axis for the three graphs shows the copy number (CN) status determined from the array for a variant at exon 9 only (left), for the overall CN status at a probability threshold of 0.90 (middle), and for the overall CN status at a probability threshold of 0.97 (right). b) the CN status for the four deviating samples with a probability threshold of 0.90 over all variants included in the CNV-calling algorithm. c) Haplotype frequency differences compared to the frequencies reported by CPIC (left) and contraindication frequency differences compared to the frequencies reported by CPIC (right).

Based on the validated variants, final translation tables were created to first translate variants to haplotypes (Tables S3a-x) and then to translate diplotypes to CI (Tables S4a-l). These tables were then used as input for *Asterix* to create preliminary PGx profiles for 996 Lifelines participants and 470 Lifelines NEXT participants. Approval (or adjustment) and release by a registered pharmacist resulted in the final PGx profiles. Haplotype frequencies were compared to reference frequencies in a European population (source: Clinical Pharmacogenetics Implementation Consortium (CPIC)) or NFE (source: DPWG/gnomAD) (Figure 4 and Tables S6a-b).

Finally, the technical PGx profiles were shared with the biobank participants in the smartphone application *Gen en Geneesmiddel* (Figure 5). The app links the technical information in the PGx profile with associated explanatory texts, yielding a layperson-friendly PGx passport. Rather than using genes as the basic unit of information, the app categorizes drugs into five categories (Figure 5a): poor metabolism (‘very slow’), intermediate metabolism (‘slow’), normal metabolism (‘average’), ultrarapid metabolism (‘very fast’), and non-metabolic drug–gene interactions (‘other processes’). By selecting one of the categories, users can see a list of all the medications in that category, based on their pharmacogenetic profile (Figure 5b). More information can be found on specific medication (Figure 5c), i.e., what is it used for and how is it affected by the user’s PGx profile. Interested users can get more information on the associated CIs by proceeding to a linked article. Finally, the technical information (PGx profile) can be downloaded and/or shared directly from the app (Figure 5d).

**Figure 5:**
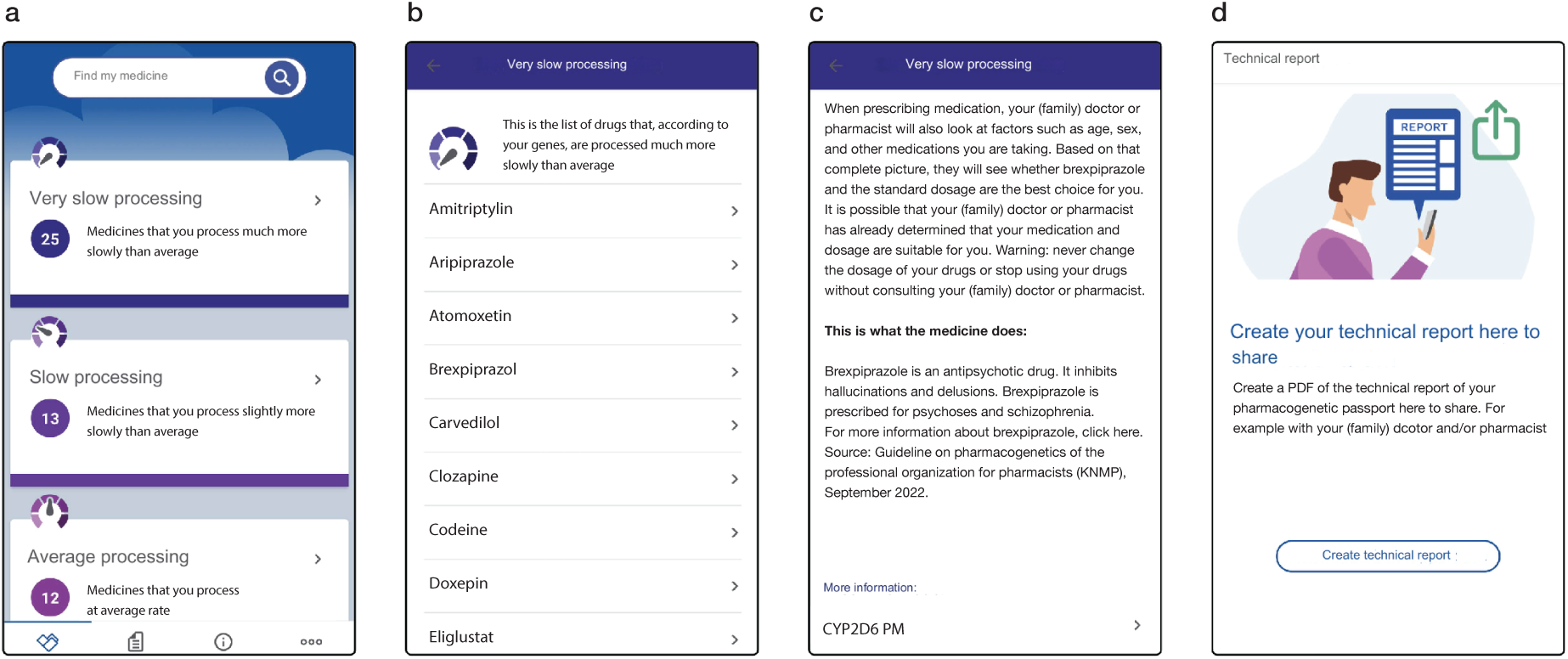
Overview of the ‘Gen en Geneesmiddel’ app (translated from Dutch). a) Main menu, where five categories can be selected: ‘poor’, ‘intermediate’, ‘normal’ and ‘ultrarapid’ metabolism and ‘other processes’. b) A personalized list of medication in the ‘poor metabolism’ category. c) Explanation of the predicted metabolism of brexpiprazole. d) The pharmacogenetic profile can be saved or shared from the app.

Based on the PGx profiles we generated using *Asterix*, 92% and 90% of the Lifelines and Lifelines NEXT populations, respectively, carried at least one actionable PGx CI (Tables S6a-b list frequencies of PGx haplotypes and CIs). We have returned these profiles to the biobank participants through the *Gen en Geneesmiddel* app, enabling them to get optimized (future) drug prescriptions with the help of their physician and pharmacist.

## 4. Discussion

In this study, we use oligonucleotide genotyping arrays to develop a low-cost PGx passport that repurposes existing genotype data from the Lifelines and Lifelines NEXT cohorts for clinical use. We clinically validated the genotyping method by aligning validation parameters and corresponding acceptance criteria to the local clinical diagnostic center and by comparing genotyping results with results from classical PCR-based assays. Using parent–offspring trios, we could also verify the accuracy of haplotype phasing, information that is unavailable when using PCR-based PGx assays. We showed that we could confidently include most of the recommended ‘tier 1’ PGx variants for the genes included in our study (26–29), with only a few exceptions. *TPMT* was excluded for the GSA-MDv3 oligonucleotide array because variant rs1800462 did not meet validation criteria. In addition, three *CYP2C9* variants classified as tier 1 (mutant allele frequency > 0.001) were excluded from the crucial backbone on the grounds of their much lower allele frequencies in the European population. To apply our pipeline in a broader context, these variants would need to be validated in a population with higher MAFs.

Furthermore, to address previously identified barriers to use of PGx passports by laypeople, we implemented communication of relevant PGx information through a newly developed smartphone application. As the uptake of the PGx passport increases, support will be provided for healthcare professionals to properly apply PGx in their daily practice.

### 4.1 Producing PGx profiles

As the generation of clinically valid PGx profiles from array-based genotype data is the cornerstone of this study, we discuss this first. We did not include *DPYD, FV, HLA-A, HLA-B, CYP1A2,* and *COMT* in the PGx passport because: 1) *DPYD* genotyping is already pre-emptively performed in combination with phenotyping in routine care, 2) the DPWG guidelines for mutations in *FV* were retracted, 3) there is no allele definition table for determining *HLA* alleles, so we could not use the same pipeline for this region, and 4) *CYP1A2* and *COMT* are part of DPWG guidelines but currently do not contain DGIs.

An important limitation in our passport production is that we could not provide a complete passport for every individual. The reason is twofold: First, we used imputation quality (R^2^) to select imputed variants of sufficient quality. If a variant with insufficient imputation quality (R^2^ < 0.97) was in the crucial backbone, the variant was not removed from the translation tables, as that would lead to the complete exclusion of the corresponding gene for all individuals. Instead, we use haploid dosage scores to select a subset of individuals with imputed genotypes of sufficient quality. We could thereby still report results for a subset of individuals. When applying our method in a diagnostic setting, genotypes should be determined by a back-up analysis if no reliable result can be extracted from the array data. Second, our *CYP2D6* CNV-calling is currently aimed at detecting whole gene deletions and duplications, ignoring potential hybrid alleles. While this is an important limitation, our method is already able to determine CNV-status on the level of single variants as opposed to the full locus. So far, we have used this to our advantage to ensure that CNV-status across the entire gene is consistent, before a whole gene deletions or duplications is called. However, this could also be used to call hybrid alleles, allowing us to assess structural variation on a much higher resolution than currently achieved in diagnostic settings, without the need for more expensive sequencing technologies. Within the scope of this study however, we were unable to develop this.

We confirm that actionable PGx variants are present in >90% of the general population (Table S7), comparable to the percentages reported in literature. In this implementation study, we were unable to evaluate the impact of these actionable variants on drug efficacy and adverse effects, due to the scale of our study and the absence of high-quality prescription records for our study populations. In the future, we aim to further investigate this by linking prescription data from an external database through the PharmLines initiative (28).

### 4.2 Layperson-friendly communication of PGx passports

Engaging patients with PGx testing is not limited to increasing their understanding of genetic information, it is also meant to enable their participation in prescription decisions. Patients who are well-informed about the implications for their prescription management are better equipped to collaborate with their healthcare providers in developing personalized treatment plans that align with their needs and preferences (32). Furthermore, this engagement builds a sense of empowerment and ownership over one’s health, which has been shown to improve adherence to treatment regimens and to lead to better health outcomes (33).

The PGx passport, as a digital health tool, facilitates patient engagement by providing a platform to exchange PGx information and medication management to enhance patient understanding. By providing layperson-friendly communication within the PGx passport, it may in the future be used to support patient education initiatives to broaden the understanding and implementation of pharmacogenomics across the Netherlands.

We aimed to develop communication materials that are understandable for 80% of the general population. Since the Lifelines and Lifelines NEXT cohorts are more highly educated than the general population, we speculate that the material should be understandable for around 90% of the study population. In an ongoing implementation study, we will evaluate the level of comprehension among participants, so that texts can be better adapted to citizens’ needs.

Several practical decisions were made in the development of the smartphone application. For example, *CYP3A5* non-expressers (*3/*3) were placed in the app category that represents normal metabolism, while both hetero- and homozygous expressers are placed in the ultrarapid metabolism category. In the Netherlands, *CYP3A5* non-expressers are most prevalent, so the standard of care applies for people with this metabolizer status.

### 4.3 Future perspectives

The completion of the PGx passport and smartphone application represents another step forward on the road toward true personalized medicine. By demonstrating to healthcare professionals how incorporating PGx into daily practice works to streamline prescribing practices through the added value of reduced adverse effects, the PGx passport aims to lower the barrier to making key prescription decisions.

Although clinical implementation of PGx has picked up pace in the past years, it has not yet been integrated into routine care. One of the main barriers here is likely to be the lack of strong evidence that its application effectively improves healthcare and reduces healthcare costs, which are both prerequisites for reimbursement (34).

Other important potential barriers are the complexities associated with integrating genetic data into electronic health records and the need to ensure that healthcare professionals have the requisite expertise and resources to comprehend and utilize PGx insights (35,36). Previous research at our center has shown that, although these barriers exist, they can be overcome (2).

In this study, we have demonstrated that PGx passports can be generated much more cheaply than the currently prevailing methodologies. However, the efficacy and cost-effectiveness of pre-emptive PGx testing still needs to be more thoroughly proven, and this will require the inclusion of large numbers of participants from the general population. Only then will the numbers of drug prescriptions dispensed based on the information in a PGx passport be sufficient to perform quantitative analysis of adverse effects and/or treatment effectiveness. As the PGx passport is currently limited to the Netherlands, the findings on cost-effectiveness would only be applicable to the Netherlands. However, with the push to harmonize the European Health Data Space, demonstrating cost-effectiveness in the Netherlands will enrich the changing digital health space. Lastly, as the initial data was derived from the Lifelines biobank, incorporating *Asterix* into future investigations in Lifelines would enable more in-depth research into the interplay between genetic variation, environment, population health, and pharmacotherapy.

### 4.4 Concluding remarks

This study provides a method to produce low-cost, clinically validated PGx profiles for use by healthcare professionals from genotype array data. As more and more individuals are genotyped, both in academic and healthcare settings, translating available genetic data towards clinical decision-making will be essential for personalized care. Furthermore, we have developed a smartphone application to ensure that the results from our pipeline, *Asterix,* reach those whose health may be impacted. To break down barriers to patient engagement, the application provides a novel approach to understandably communicate PGx results to laypeople. The reception of this information by users will be reported in an upcoming publication.

Future steps include the evaluation of the costs and effectiveness of the pre-emptive PGx passport in the general population and further research into the interplay between genetic variation and population health and drug response phenotypes.

## Acknowledgements

The authors thank the UMCG Genomics Coordination Center, the UG Center for Information Technology and their sponsors BBMRI-NL & TarGet for storage and compute infrastructure. They thank Maaike van der Lee and Jesse Swen from the department of Clinical Pharmacy and Toxicology of the Leiden University Medical Center for providing the validation samples with known genotypes. The authors thank Certe Groningen for their assistance in genotyping the CYP2D6 gene in a number of cases and the GIC of the KNMP for providing their drug information texts to generate layperson-friendly information. They also thank past and present members of the ELSI-team of the UMCG for valuable feedback on the developed communication materials.

## Lifelines Cohort Study

The Lifelines Biobank initiative has been made possible by funding from the Dutch Ministry of Health, Welfare and Sport, the Dutch Ministry of Economic Affairs, the University Medical Center Groningen (UMCG the Netherlands), University of Groningen and the Northern Provinces of the Netherlands. The generation and management of GWAS genotype data for the Lifelines Cohort Study is supported by the UMCG Genetics Lifelines Initiative (UGLI). The authors wish to acknowledge the services of the Lifelines Cohort Study, the contributing research centers delivering data to Lifelines, and all the study participants.

Raul Aguirre-Gamboa (1), Patrick Deelen (1), Lude Franke (1), Jan A Kuivenhoven (2), Esteban A Lopera Maya (1), Ilja M Nolte (3), Serena Sanna (1), Harold Snieder (3), Morris A Swertz (1), Judith M Vonk (3), Cisca Wijmenga (1)

(1) Department of Genetics, University of Groningen, University Medical Center Groningen, The Netherlands

(2) Department of Pediatrics, University of Groningen, University Medical Center Groningen, The Netherlands

(3) Department of Epidemiology, University of Groningen, University Medical Center Groningen, The Netherlands

## Author contributions

PL, BW, MS, NK, JD and LF: conceptualization,

PL, RW, HB, TO, JS, MB, ELM, TOM, PD, JD and LF: methodology,

RW, HB, BC, JS, LJ and LF: software, PL, RW, HB, TO, JS and MB: validation,

PL, RW, HB, TO, JS, MB, ELM and SJ: formal analysis, SJ, JGA and DTe: investigation,

LLN, LLCS, PD, NK, JD and LF: resources,

PL, RW, HB, JS, ELM, SJ, BC and LJ: data curation,

PL, RW, HB, TO and JS: manuscript – original draft, RW and JS: visualization,

BW, DTo, PD, NK, JD and LF: supervision, PL, JS and JD: project administration,

NK, JD and LF: funding acquisition.

## Data availability

Due to privacy reasons the individual-level data cannot be made publicly available but can be made available upon reasonable request. The individual-level data from Lifelines that support the findings in this publication were obtained from the Lifelines biobank under project application number ov21_0355. This request should be directed to the Lifelines Research Office through email or by using the application form on their website (https://www.lifelines.nl/researcher/how-to-apply/apply-here). Access to the Lifelines NEXT Project data will be granted to all qualified researchers and will be governed by the provisions laid out in the LLNEXT Data Access Agreement: https://groningenmicrobiome.org/?page_id=2598. This access procedure is in place to ensure that the data is requested solely for research and scientific purposes, in compliance with the informed consent signed by Lifelines NEXT participants. Scripts and other software central to this manuscript is available on GitHub (https://github.com/molgenis/PGx-passport-pilot/ and https://github.com/molgenis/asterix). Other analyses of genotype data and publicly available reference data is performed using standard bioinformatics practices, for which the code is made available upon request.

