## Supplemental Information for "Low-cost generation of clinical-grade layperson-friendly pharmacogenetic passports using oligonucleotide arrays"

**Supplementary methods**

*Approval and release of PGx profiles*

After the automated generation, by Asterix, of a preliminary PGx profile, profiles needed to be approved and released by a registered pharmacist. To support the pharmacist in this process, the following parameters from Asterix output were used: *detected mutations* in the gene under consideration; *low-quality calls* ($p < 0.90$); *intermediate-quality calls* ($0.90 \leq p < 0.97$); the *haplotype call made by Asterix*, including the *associated disclaimer*; and, for *CYP2D6*, *CNV-status* and its confidence level. If the haplotype to be approved contained a combination of mutations and/or low- or intermediate-quality calls that had never been approved previously, the pharmacist was presented with all the relevant information and was required to either approve the call made by Asterix (and the associated disclaimer), or to input a corrected call and disclaimer.

If a variant from the *crucial backbone* was called with insufficient confidence level (< 0.97), the haplotype (and by extension, the gene) would be set to 'NA' (not available). Two exceptions to this rule were made: i) when the context of other detected mutations would render the call of the variant in question irrelevant (e.g. when a separate mutation was detected that leads to a premature stop codon or frameshift), ii) when the context of other detected mutations already led to a change in enzyme activity in the same direction as the variant in question. In the latter case, excluding the gene from the profile would lead to the application of standard-of-care, which was considered undesirable. A specific disclaimer was included to indicate that the presence or absence of the low-quality *crucial backbone* variant could not be excluded.

If a combination of mutations was found that did not appear in the PharmVar reference tables, the pharmacist was required to make a decision themselves. In many cases, only one of the detected mutations was documented to have an effect on enzyme activity, while other detected mutations were found in sub-alleles of the wild-type gene (*1). In that case, the star allele associated with the affecting variant was called. In some cases, multiple variants were detected with either conflicting or unknown effect on enzyme activity. In those cases, the haplotype would be set to 'NA'.

The decision made was then logged, both to keep a clear audit trail, but also to allow re-application of the decisions made: if the haplotype to be approved contained a previously approved combination of mutations and/or low- or intermediate-quality calls, the decision and disclaimer from the decision logging was automatically adopted without intervention by the pharmacist.

While approval of other genes was only performed on haplotype-level, copy number variation in *CYP2D6* necessitated an additional step for approval. Firstly, *CYP2D6* was set to 'NA' if i) no confident CNV-call was available, or ii) more than 2 copies were reliably detected (as imputation and phasing were lacking for those samples). Secondly, if 0 copies were detected, the gene was directly called as '*5/*5'. If 2 copies were detected, *CYP2D6* approval was performed as for other genes (see above). Finally, if 1 copy of *CYP2D6* was detected, all variants that were reanalyzed in the CNV-calling procedure were written as homozygous in the genotype file. Subsequently, *CYP2D6* was imputed and phased as if 2 copies were present. If Asterix then assigned the same star allele (*x) to both haplotypes, the gene was called as '*5/*x'. If Asterix assigned two different star alleles to the two haplotypes in the genotype data, *CYP2D6* was set to 'NA', as we currently cannot properly phase whole-gene deletions with the surroundings of the gene.
